# Shotgun metagenomic sequencing analysis as a diagnostic strategy for patients with lower respiratory tract infections

**DOI:** 10.1101/2025.04.24.25326335

**Authors:** Ha-eun Cho, Min Jin Kim, Jongmun Choi, Yong-Hak Sohn, Jae Joon Lee, Kyung Sun Park, Sun Young Cho, Ki-Ho Park, Young Jin Kim

**Affiliations:** Department of Laboratory Medicine, Kyung Hee University Medical Center, Seoul, Republic of Korea; Department of Laboratory Medicine, Kyung Hee University School of Medicine, Seoul, Republic of Korea; Department of Laboratory Medicine, Seegene Medical Foundation, Seoul, Republic of Korea; Division of Infectious Diseases, Department of Internal Medicine, Kyung Hee University Medical Center, Seoul, Republic of Korea; Division of Infectious Diseases, Department of Internal Medicine, Kyung Hee University School of Medicine, Seoul, Republic of Korea

## Abstract

**Introduction:** Diagnosing lower respiratory infections (LRIs) using conventional diagnostic methods (CDMs) presents limitations in detecting suspected pathogens. This study compares the latest CDMs with shotgun metagenomic sequencing (SMS) for bronchoalveolar lavage (BAL) fluid. The primary objective is to enhance pathogen detection using SMS.

**Materials and Methods:** A total of 16 BAL fluid samples from patients with pneumonia with positive results in various CDMs—bacterial/fungal cultures, real-time PCR for *Mycobacterium tuberculosis*, cytomegalovirus, or the BioFire® FilmArray Pneumonia Panel—were included. Samples were subjected to 10 Gb SMS on the NovaSeq 6000 (Illumina) and were aligned against the NCBI RefSeq database. For eukaryotic reads, an additional matching process was performed using the internal transcribed spacer (ITS) region of fungi. Antibiotic resistance genes (ARGs) were annotated using the Comprehensive Antibiotic Resistance Database model. To identify significant pathogens, thresholds for the relative abundance of SMS reads were applied to evaluate the concordance between CDM- and SMS-detected microbes.

**Results:** The proportion of microbial reads ranged from 0.00002–0.04971% per sample. SMS detected corresponding bacterial reads (2–23,869) with relative abundance between 0.02% and 87.5%. Eukaryotic reads varied from 0 to 32, with no fungal alignment at the genus level. *Candida* species were identified in four samples using ITS. No viral reads were detected. In 10 out of 16 cases (63%), SMS detected pathogens above the threshold by SMS. When subdominant taxa were included, SMS detected pathogens in 11 out of 16 cases (69%). ARGs meeting perfect criteria via the Resistance Gene Identifier were observed in two cases.

**Conclusion:** This study represents the first comparison of SMS and CDMs, including the FilmArray Pneumonia Panel, in the context of LRI diagnostics. SMS may serve as a valuable supplementary tool for LRI diagnosis. Further research is necessary to improve sensitivity and cost-effectiveness.

## Introduction

Identifying the causative pathogen of infectious pneumonia is crucial for targeted treatment and improved patient outcomes (1). The detection rate of pathogens in patients with lower respiratory infections (LRIs) ranges from 38–46% (2, 3). Despite the identification of multiple microorganisms using conventional diagnostic methods (CDMs), a single pathogen is typically considered the primary cause of infection (4). Although polymicrobial infections are reported in 5.7–38.4% of LRI cases, their etiology is rarely confirmed during treatment (5–9).

Bacterial etiology accounts for over 50% of diagnoses, leading to the prioritization of empirical antibiotic use (10). However, the spectrum of causative microbes is diverse (11). Culture and polymerase chain reaction (PCR) are standard diagnostic tools supplemented by antigen tests. However, cultures may fail to detect fastidious bacteria and fungi (12, 13), while targeted PCR may overlook microbes not included in the assay (14). Multiplex PCR panels expand the detection range of clinically relevant microbes, enabling comprehensive identification. Nevertheless, these PCR panels have limitations, particularly when pathogens not covered in the panel proliferate and contribute to infection (15). Antigen tests provide a rapid and cost-effective diagnostic option but are hindered by low sensitivity and false positives (13). The variability in pathogen detection rates across these diagnostic methods highlights the complexity of achieving accurate LRI diagnoses, emphasizing the need for complementary diagnostic approaches.

In response, shotgun metagenomic sequencing (SMS) has gained attention. This approach involves analyzing all nucleic acids within a sample, enabling the comprehensive identification of potential pathogens. Furthermore, the use of extensive reference databases enhances the potential for syndromic testing, offering universal pathogen detection (16). Studies utilizing SMS on bronchoalveolar lavage (BAL) specimens have demonstrated a sensitivity of 88–97% and a specificity of 15–81% for accurate pathogen identification (17–20).

Owing to its anatomical location, BAL fluid inherently possesses low microbial biomass (21), posing challenges for microbial signal detection. Therefore, optimizing methodologies is crucial for achieving reliable SMS results. Despite the development of semiquantitative multiplex PCR, few studies have compared its performance to that of SMS. Furthermore, discussions on the appropriate capacity for SMS of BAL fluid remain limited. The absence of consensus on the criteria for interpreting SMS positivity further complicates the issue (17–20, 22–26).

This study presents the first comparative assessment of SMS and conventional diagnostic tests in clinical practice, including semiquantitative multiplex PCR. The performance of SMS with a 10 Gb output was evaluated, along with efforts to optimize the criteria for SMS interpretation.

## Materials and Methods

### Sample collection and processing

From March to July 2023, a total of 44 BAL fluid samples with positive results were consecutively collected from the BioFire® FilmArray® Pneumonia Panel (FA-PP; BioFire Diagnostics LLC, Salt Lake City, UT, USA). The FA-PP is a semi-quantitative PCR test representing the latest diagnostic method for pathogen detection. Exclusion criteria for sample selection were as follows: 1) clinical diagnosis of non-infectious pneumonia, 2) contamination with normal flora (27), and 3) final diagnosis of pneumonia caused by RNA viruses. Initially, 12 cases diagnosed with non-infectious diseases were excluded, leaving 32 cases for further analysis (Fig 1).

**Fig 1.**
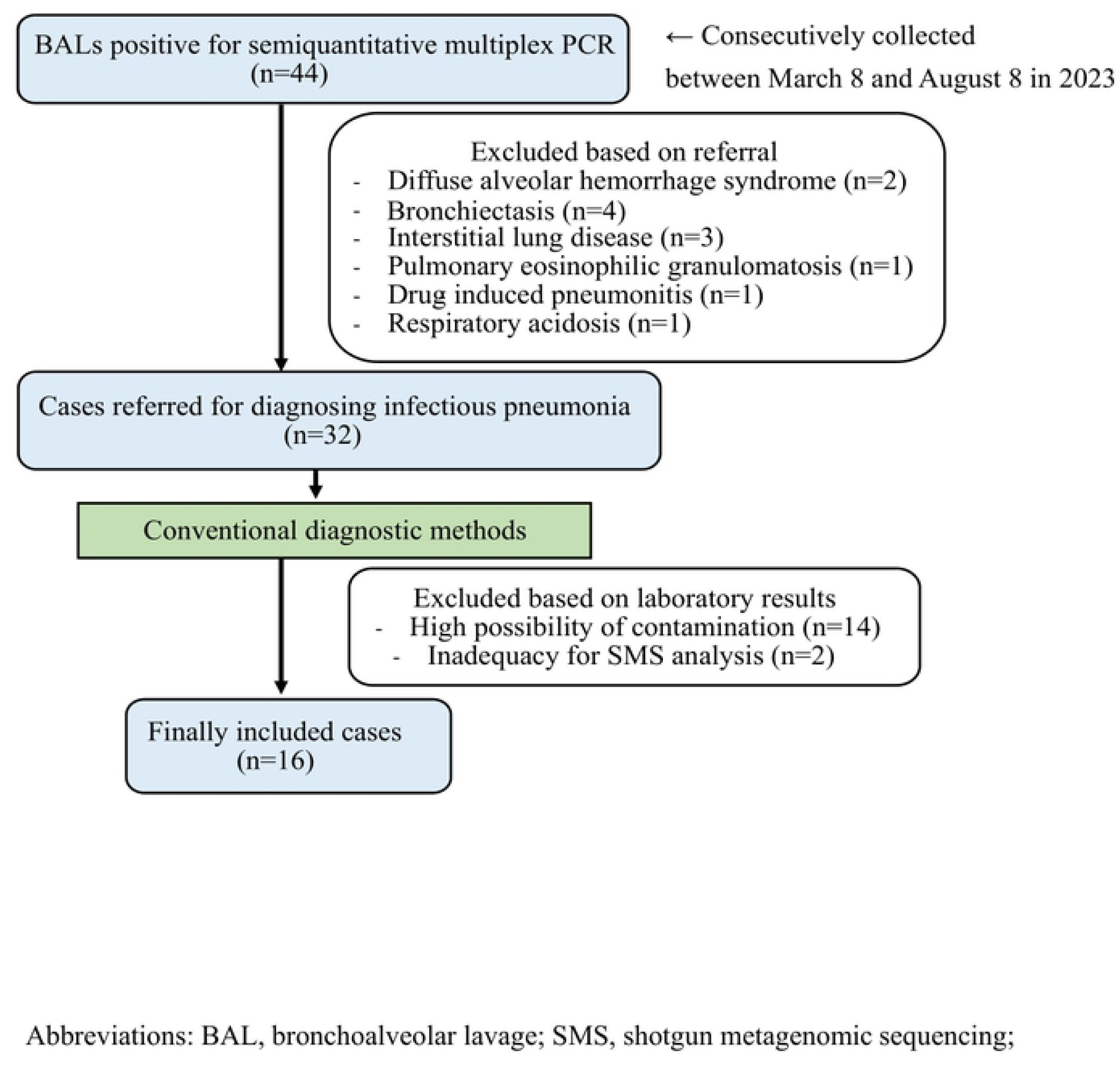
Flow diagram of case inclusion and exclusion.

BAL fluids with positive FA-PP results were consecutively collected over a 5-month period. Among these, 16 cases were selected for SMS analysis after excluding samples with high contamination risk or those with CDM results detecting only RNA viruses.

CDMs were also performed. In this study, CDMs referred to currently used clinical methods for pathogen detection, including bacterial, fungal, and *Mycobacterium tuberculosis*/non-tuberculous mycobacteria (MTB/NTM) cultures. Additional tests included matrix-assisted laser desorption/ionization time-of-flight mass spectrometry system (Bruker Daltonics, Billerica, MA, USA), TB/NTM PCR (AdvanSure™ TB/NTM real-time PCR kit on AdvanSure SLAN 96 and E3 system; LG Life Science, Seoul, Korea), Xpert MTB/RIF assay (Cepheid, Sunnyvale, CA, USA), cytomegalovirus (CMV) PCR (nucleic acid extracted using QiaAmp DSP DNA mini kit on QIAcube; Qiagen GmbH, Hilden, Germany and PCR performed on CFX 96; Bio-Rad, Hercules, CA, USA), *Pneumocystis jirovecii* PCR (nucleic acid extracted using a laboratory-developed test reagent on MagNa Pure 96; Roche Diagnostics, Mannheim, Germany, and PCR performed on CFX 96; Bio-Rad, Hercules, CA, USA), *Aspergillus* antigen test (PLATELIA Aspergillus Ag; Bio-Rad, Hercules, CA, USA), and *Cryptococcus* antigen test (Pastorex Crypto Plus; Sanofi-Diagnostics Pasteur, Marnes-La-Coquette, France). The positive reporting criterion for microbial growth was defined as greater than 10^4^ CFU/mL in culture (27). FA-PP positivity was reported when it exceeded 10^4^ copies/mL.

Based on culture results, 14 samples with a high risk of contamination were additionally excluded. These included samples containing normal oropharyngeal flora, which may have been introduced during the BAL procedure (28), and normal cutaneous flora, which could have led to contamination during specimen collection and processing (29). However, commensal microorganisms with a high potential to cause pneumonia, such as *Staphylococcus aureus*, as well as cases in which pathogens such as TB were co-detected, were included in the study (30). Given that this study utilized DNA-based SMS and did not target RNA, two additional samples in which only RNA viruses were identified via CDMs were excluded. Ultimately, 16 selected samples were included in this study.

### Extraction of nucleic acids and library preparation

DNA was extracted from 16 BAL fluid samples using the QIAamp DNA Mini Kit (Qiagen, Hilden, Germany). The extracted nucleic acid concentration was measured using the Qubit dsDNA HS Assay Kit on a Qubit 4.0 instrument (Life Technologies Carlsbad, CA, USA) and the Nanodrop One (Thermo Fisher Scientific, Waltham, MA, USA). DNA integrity was assessed using the Genomic DNA ScreenTape Assay Kit on a TapeStation 4200 System (Agilent Technologies, Inc., Santa Clara, CA, USA).

Libraries were constructed using Illumina DNA Prep (Illumina, San Diego, CA, USA). Quality control was performed using the TapeStation 4200 System with the D1000 ScreenTape Assay Kit (Agilent Technologies, Inc.).

### SMS procedures

Shotgun sequencing was performed using a paired-end configuration with 150 bp × 2 reads on the NovaSeq 6000 instrument (Illumina, San Diego, CA, USA), utilizing SP Flow Cell reagents. The sequencing output was set to a capacity of 10 Gb. To assess the quality of the generated raw data, FastQC software (version 0.12.0) was used to examine the proportion of reads with a quality score of Q30 or higher and the GC content.

Adapter sequences were removed through trimming. Raw reads were mapped to the human host reference genome (GRCh38.p14) using URMAP and Samtools. Unmapped reads were extracted and merged using the BBMap software. MMseq2 was employed to align sequences against the SILVA_138 database. Initially, an e-value cutoff of 1e-5 was applied to identify and intersect unique genes in each ecosystem. For taxonomic assignment, specific parameters included a minimum alignment length of 90 and an identity threshold of 80, with the highest values prioritized. All unmapped reads were assembled using SPAdes, applying the metaviral option to isolate scaffolds corresponding to viral genomes. Reference DNA viruses were selected based on their relevance to LRIs, including adenovirus, CMV, Epstein-Barr virus, varicella-zoster virus, and herpes simplex virus (3, 11, 31, 32). Databases were downloaded from the National Center for Biotechnology Information (NCBI) RefSeq (version 2023.10.13) for classification. Bacterial sequences that could not be classified at the genus or species level were designated as unclassified bacteria. Fungal analysis was conducted based on the internal transcribed spacer (ITS) gene for fungi confirmed by fungal culture and PCR. To profile the resistome, the Comprehensive Antibiotic Resistance Database (CARD) Resistance Gene Identifier (RGI) software (version 6.0.3) and the CARD database (version 3.2.8) were used, assigning genes to the CARD model for annotation of antibiotic resistance genes (ARGs). The raw sequencing data were submitted to the NCBI Sequence Read Archive (SRA) database under the accession number PRJNA-1036216.

The criteria for pathogen detection in SMS were applied based on existing literature. For bacterial detection, a relative abundance threshold of ≥30% was applied (17, 18, 20, 26). TB was considered positive with ≥1 mapped read (17–19, 22–24, 26), while NTM were classified as positive if they were among the top 10 bacterial taxa (23, 24). For fungal detection, a species was considered positive if its coverage rate was at least five times higher than that of any other fungal species (22, 23, 26). However, when the coverage rate was insufficient for positive determination, fungal reads were considered positive if they met the following criteria: an alignment length ≥100 bp and an identity of ≥98%. Viral detection required a minimum of three mapped reads (18, 24, 33).

Microbial reads that met these thresholds were classified as SMS-positive. Subdominant microbial reads that did not meet these thresholds were also reviewed. In each case, the microbe determined by the attending physician to be the causative agent of the infection and targeted for antimicrobial therapy was considered clinically relevant.

The identified ARGs were determined using the "perfect" algorithm of the RGI, which detects exact matches to curated reference sequences and known resistance-conferring mutations (34). ARG results were compared with the antibiotic susceptibility test (AST) results of cultured strains from the samples. If the cultured strain exhibited resistance to antibiotics associated with the detected ARGs, the result was classified as consistent. Conversely, if any antibiotic within the corresponding group showed susceptibility or an indeterminate result in AST, the finding was classified as inconsistent.

### Ethics statement

Ethical approval for this study was obtained from the Institutional Review Board of Kyung Hee University Medical Center (no. 2023-05-076) on June 1, 2023. The approved collection period for archived residual samples extended until February 29, 2024. As only archived samples were used and all data were fully anonymized prior to analysis, the requirement for informed consent was waived by the IRB.

## Results

### Identification of microbes by SMS

Among the 16 FA-PP-positive samples, microbial reads were detected in all cases, accounting for 0.00002–0.04971% of the total reads. Of these, 99.3% were bacterial reads matched to the 16S rRNA gene, 0.3% were eukaryotic reads matched to the 18S rRNA gene, and 0.4% were fungal reads matched to the ITS gene.

SMS detected pathogens above the threshold in 10 of 16 cases (63%), increasing to 11 of 16 cases (69%) when subdominant taxa were included. CDMs identified bacterial pathogens in 11 of 16 cases (69%), including two cases of MTB. Among the 16 cases, bacterial–fungal co-infections were observed in 2 of 16 cases (12.5%), a bacterial–viral co-infection in 1 case (6.25%), and a fungal–viral co-infection in 1 case (6.25%). In the 11 cases in which CDMs identified bacteria as the causative pathogen, SMS detected the corresponding bacteria in nine cases. These included *Pseudomonas aeruginosa* in cases 3, 12, 14, and 15; *Haemophilus influenzae* in cases 10 and 11; *Klebsiella pneumoniae* in case 7; *Acinetobacter baumannii* in case 13; and *Stenotrophomonas maltophilia* in case 16. Expanding the criteria to include subdominant bacterial reads, *K. pneumoniae* was additionally detected in case 8, increasing the detection rate to 10 out of 11 bacterial cases. In cases where fungi were the primary pathogen, SMS detected *Candida tropicalis* reads in case 4 (Table 1).

**Table 1.**
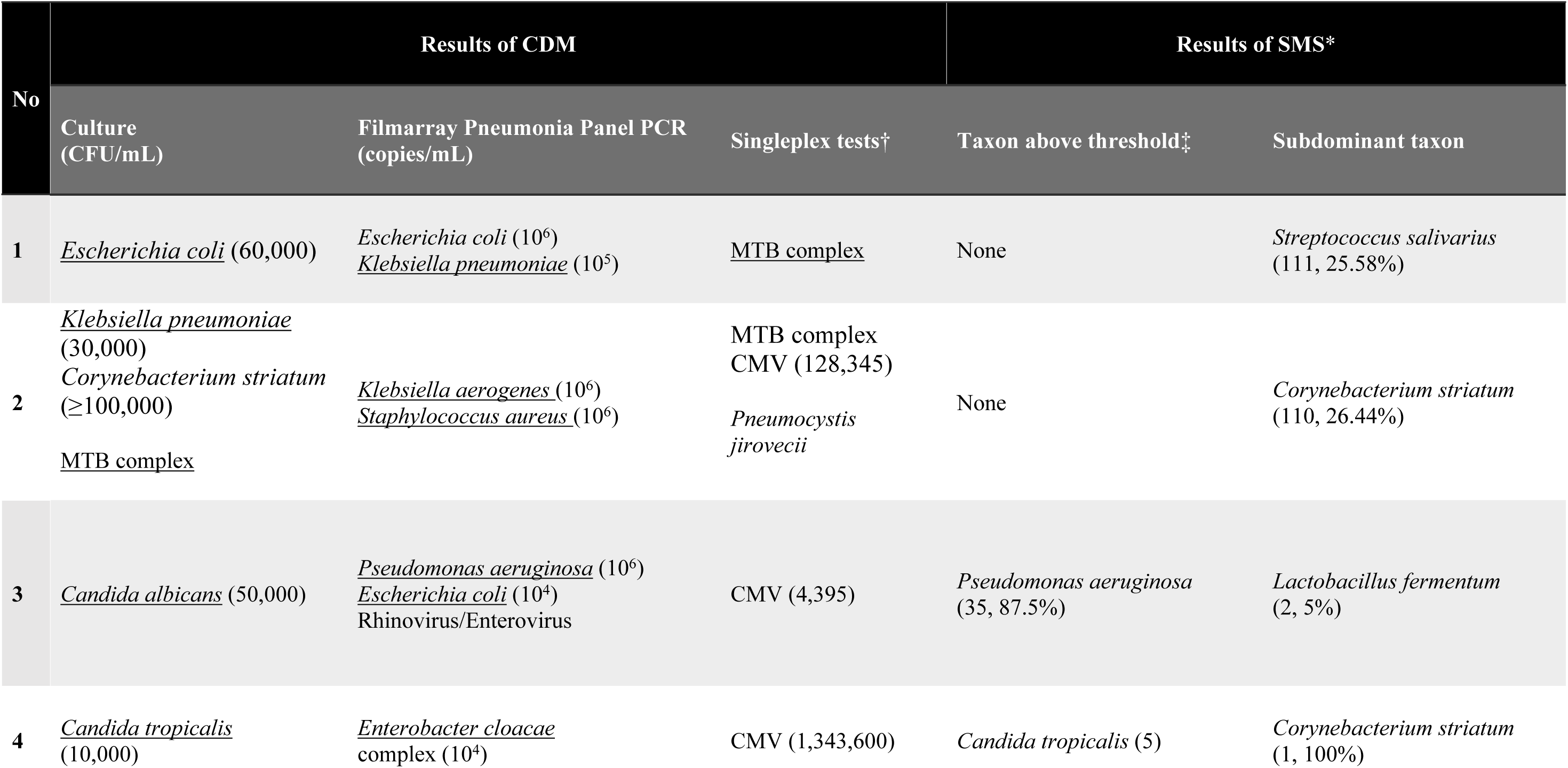
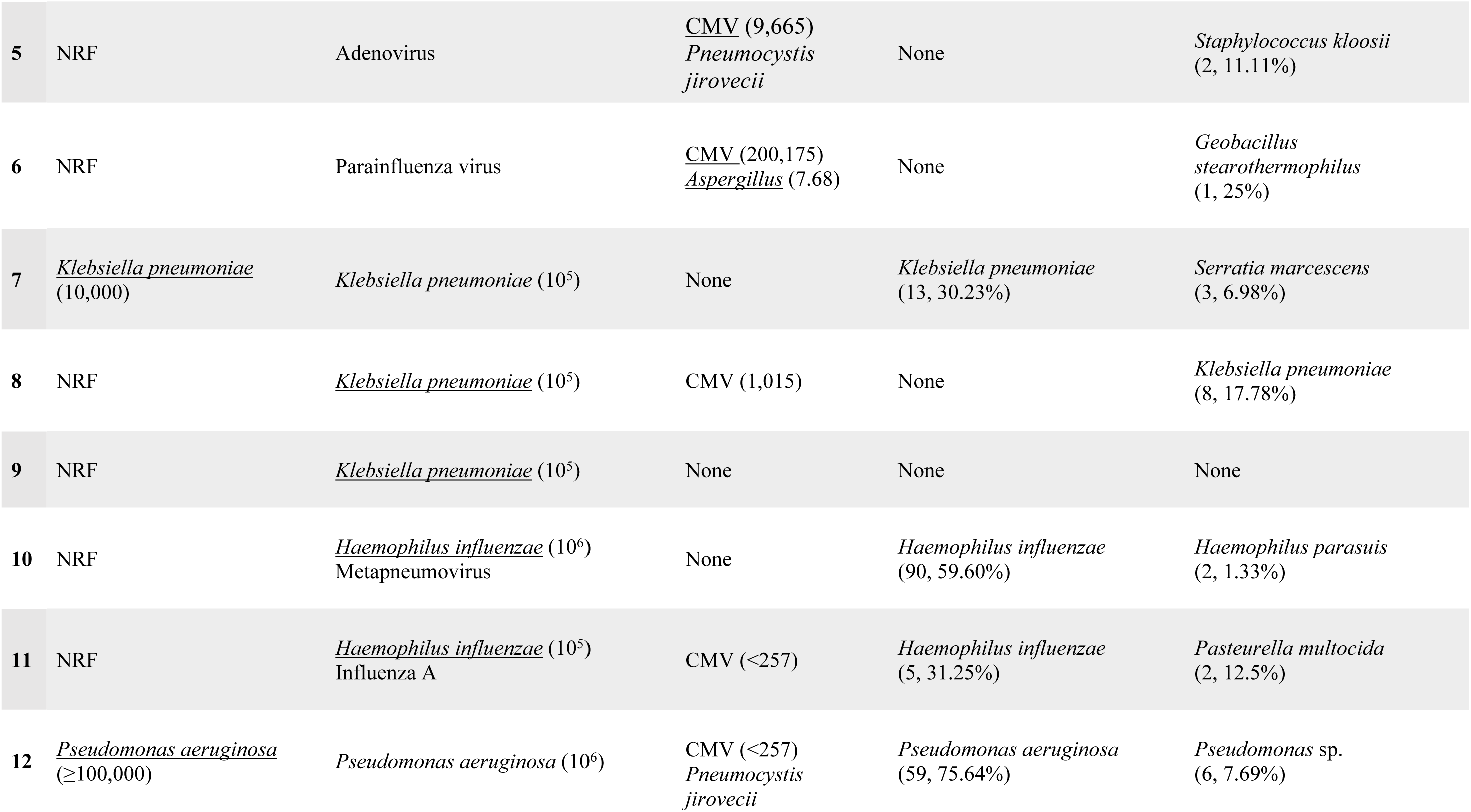
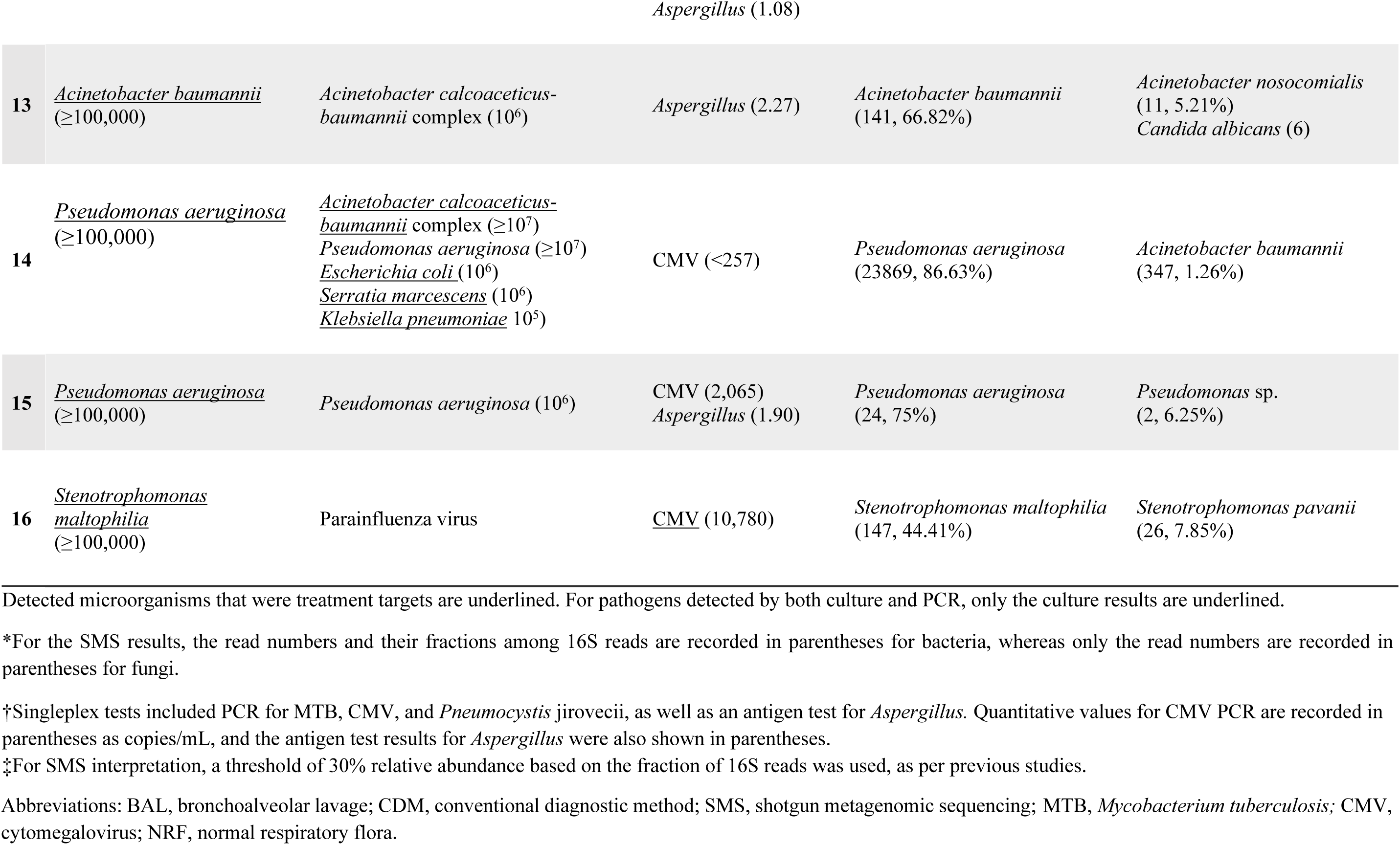
Comparison of microbial results from CDMs and SMS.

| No | Results of CDM |  |  | Results of SMS* |  |
| --- | --- | --- | --- | --- | --- |
|  | Culture (CFU/mL) | Filmarray Pneumonia Panel PCR (copies/mL) | Singleplex tests† | Taxon above threshold‡ | Subdominant taxon |
| 1 | *Escherichia coli* (60,000) | *Escherichia coli* (10<sup>6</sup>)<br>*Klebsiella pneumoniae* (10<sup>5</sup>) | MTB complex | None | *Streptococcus salivarius* (111, 25.58%) |
| 2 | *Klebsiella pneumoniae* (30,000)<br>*Corynebacterium striatum* (≥100,000)<br>MTB complex | *Klebsiella aerogenes* (10<sup>6</sup>)<br>*Staphylococcus aureus* (10<sup>6</sup>) | MTB complex<br>CMV (128,345)<br>*Pneumocystis jirovecii* | None | *Corynebacterium striatum* (110, 26.44%) |
| 3 | *Candida albicans* (50,000) | *Pseudomonas aeruginosa* (10<sup>6</sup>)<br>*Escherichia coli* (10<sup>4</sup>)<br>Rhinovirus/Enterovirus | CMV (4,395) | *Pseudomonas aeruginosa* (35, 87.5%) | *Lactobacillus fermentum* (2, 5%) |
| 4 | *Candida tropicalis* (10,000) | *Enterobacter cloacae* complex (10<sup>4</sup>) | CMV (1,343,600) | *Candida tropicalis* (5) | *Corynebacterium striatum* (1, 100%) |
| 5 | NRF | Adenovirus | CMV (9,665)<br>*Pneumocystis jirovecii* | None | *Staphylococcus kloosii*<br>(2, 11.11%) |
| 6 | NRF | Parainfluenza virus | CMV (200,175)<br>*Aspergillus* (7.68) | None | *Geobacillus stearothermophilus*<br>(1, 25%) |
| 7 | *Klebsiella pneumoniae*<br>(10,000) | *Klebsiella pneumoniae* (10<sup>5</sup>) | None | *Klebsiella pneumoniae*<br>(13, 30.23%) | *Serratia marcescens*<br>(3, 6.98%) |
| 8 | NRF | *Klebsiella pneumoniae* (10<sup>5</sup>) | CMV (1,015) | None | *Klebsiella pneumoniae*<br>(8, 17.78%) |
| 9 | NRF | *Klebsiella pneumoniae* (10<sup>5</sup>) | None | None | None |
| 10 | NRF | *Haemophilus influenzae* (10<sup>6</sup>)<br>Metapneumovirus | None | *Haemophilus influenzae*<br>(90, 59.60%) | *Haemophilus parasuis*<br>(2, 1.33%) |
| 11 | NRF | *Haemophilus influenzae* (10<sup>5</sup>)<br>Influenza A | CMV (<257) | *Haemophilus influenzae*<br>(5, 31.25%) | *Pasteurella multocida*<br>(2, 12.5%) |
| 12 | *Pseudomonas aeruginosa*<br>(≥100,000) | *Pseudomonas aeruginosa* (10<sup>6</sup>) | CMV (<257)<br>*Pneumocystis jirovecii* | *Pseudomonas aeruginosa*<br>(59, 75.64%) | *Pseudomonas* sp.<br>(6, 7.69%) |
| *Aspergillus* (1.08) |  |  |  |  |  |
| 13 | <u>*Acinetobacter baumannii*</u><br>(≥100,000) | *Acinetobacter calcoaceticus-*<br>*baumannii* complex (10<sup>6</sup>) | *Aspergillus* (2.27) | *Acinetobacter baumannii*<br>(141, 66.82%) | *Acinetobacter nosocomialis*<br>(11, 5.21%)<br>*Candida albicans* (6) |
| 14 | <u>*Pseudomonas aeruginosa*</u><br>(≥100,000) | <u>*Acinetobacter calcoaceticus-*</u><br><u>*baumannii*</u> complex (≥10<sup>7</sup>)<br>*Pseudomonas aeruginosa* (≥10<sup>7</sup>)<br><u>*Escherichia coli*</u> (10<sup>6</sup>)<br><u>*Serratia marcescens*</u> (10<sup>6</sup>)<br><u>*Klebsiella pneumoniae*</u> 10<sup>5</sup>) | CMV (<257) | *Pseudomonas aeruginosa*<br>(23869, 86.63%) | *Acinetobacter baumannii*<br>(347, 1.26%) |
| 15 | <u>*Pseudomonas aeruginosa*</u><br>(≥100,000) | *Pseudomonas aeruginosa* (10<sup>6</sup>) | CMV (2,065)<br>*Aspergillus* (1.90) | *Pseudomonas aeruginosa*<br>(24, 75%) | *Pseudomonas* sp.<br>(2, 6.25%) |
| 16 | <u>*Stenotrophomonas*</u><br><u>*maltophilia*</u><br>(≥100,000) | Parainfluenza virus | <u>CMV</u> (10,780) | *Stenotrophomonas maltophilia*<br>(147, 44.41%) | *Stenotrophomonas pavanii*<br>(26, 7.85%) |

Detected microorganisms that were treatment targets are underlined. For pathogens detected by both culture and PCR, only the culture results are underlined.
\*For the SMS results, the read numbers and their fractions among 16S reads are recorded in parentheses for bacteria, whereas only the read numbers are recorded in parentheses for fungi.
†Singleplex tests included PCR for MTB, CMV, and *Pneumocystis jirovecii*, as well as an antigen test for *Aspergillus*. Quantitative values for CMV PCR are recorded in parentheses as copies/mL, and the antigen test results for *Aspergillus* were also shown in parentheses.
‡For SMS interpretation, a threshold of 30% relative abundance based on the fraction of 16S reads was used, as per previous studies.
Abbreviations: BAL, bronchoalveolar lavage; CDM, conventional diagnostic method; SMS, shotgun metagenomic sequencing; MTB, *Mycobacterium tuberculosis*; CMV, cytomegalovirus; NRF, normal respiratory flora.

### Metagenomic results of antibiotic resistance

Results meeting the perfect criteria of the RGI were observed in cases 2 and 14.

In case 2, *K. pneumoniae* was evaluated against AST results. The *sul1* gene, associated with sulfonamide resistance, was detected and corresponded with resistance to trimethoprim/sulfamethoxazole in AST. No carbapenem-related ARGs were identified, which was consistent with AST results showing susceptibility to doripenem, ertapenem, imipenem, and meropenem. However, *AAC(6’)-Ia*, associated with aminoglycoside resistance, was detected but was inconsistent with AST results, which showed susceptibility to amikacin and gentamicin (Table 2).

**Table 2.** Antibiotic resistance genes detected in case 2 with *Klebsiella pneumoniae*.

| Antibiotic Group | Resistance Gene | Gene Detection Status | Antibiotic Susceptibility | Consistency* |
| --- | --- | --- | --- | --- |
| Carbapenem | None | Not detected | Doripenem (S), Ertapenem (S), Imipenem (S), Meropenem (S) | Consistent |
| Aminoglycoside | *AAC(6')-Ia* | Detected | Amikacin (S), Gentamicin (S) | Inconsistent |
| Sulfonamide | *SulI* | Detected | Trimethoprim/Sulfamethoxazole (R) | Consistent |

\*Antibiotic resistance genes aligned with the resistance profiles of cultured isolates.
Abbreviations: S, susceptible; R, resistant.

In case 14, *P. aeruginosa* infection was evaluated. *MexI* and *H-NS*, which are associated with fluoroquinolone and tetracycline resistance, were detected and were consistent with AST results showing resistance to levofloxacin and tetracycline. However, several inconsistencies were observed*. OXA-217, smeR, H-NS*, and *CTX-M-15*, which are associated with resistance to penicillin derivatives and cephalosporins, were detected. However, AST results showed resistance only to ampicillin, piperacillin, and cefotaxime, while susceptibility was observed for cefepime and ceftazidime. For carbapenem resistance, *AXC-1* and *OXA-217* were identified; however, AST results indicated susceptibility to doripenem, imipenem, and meropenem. Additionally, *AAC(6’)-Ia*, *cpxA*, and *smeR*, linked to aminoglycoside resistance, were detected, whereas AST results showed susceptibility to amikacin and an indeterminate result for tobramycin (Table 3).

**Table 3.** Antibiotic resistance genes detected in case 14 with *Pseudomonas aeruginosa*.

| Antibiotic Group | Resistance Gene | Gene Detection Status | Antibiotic Susceptibility | Consistency* |
| --- | --- | --- | --- | --- |
| Carbapenem | *AXC-I, OXA-217* | Detected | Doripenem (S), Imipenem (S), Meropenem (S) | Inconsistent |
| Aminoglycoside | *AAC(6')-Ia, cpxA, smeR* | Detected | Amikacin (S), Tobramycin (I) | Inconsistent |
| Fluoroquinolone and Tetracycline | *MexI, H-NS* | Detected | Levofloxacin (R), Tetracycline (R) | Consistent |
| Penicillin derivatives and Cephalosporins | *OXA-217, smeR, H-NS, CTX-M-15* | Detected | Ampicillin (R), Piperacillin (R), Cefotaxime (R), Cefepime (S), Ceftazidime (S) | Inconsistent |

\*Antibiotic resistance genes aligned with the resistance profiles of cultured isolates.
Abbreviations: S, susceptible; R, resistant; I, indeterminate.

## Discussion

The proportion of microbial reads averaged 0.00412% of the total reads, ranging from 0.00002–0.04971% per sample. This aligns with the recommended range of 0.00001–0.7% for successful SMS application in clinical settings (35). Previous studies have established absolute criteria for bacterial positivity in SMS (Table 4), typically defining a relative abundance threshold of ≥30% (17, 18, 20, 26). However, some cases exhibited SMS-detected reads for subdominant taxa that matched the primary pathogen identified by CDMs. For example, in case 8, *K. pneumoniae* was identified with a relative abundance of 17.78% but was classified as positive using adjusted criteria that considered subdominant taxa. While FA-PP reported *K. pneumoniae* at 10^5^ copies/mL, culture results indicated normal respiratory flora (NRF), which was insufficient to establish it as the primary pathogen. This finding highlights the need for caution when interpreting taxa with relative abundances below 30%, as their classification as pathogens requires additional corroborative evidence. Without clear supporting data, taxa with low relative abundances should not be automatically assumed to represent the primary pathogen. However, their potential clinical relevance warrants careful consideration depending on the clinical context.

**Table 4.**
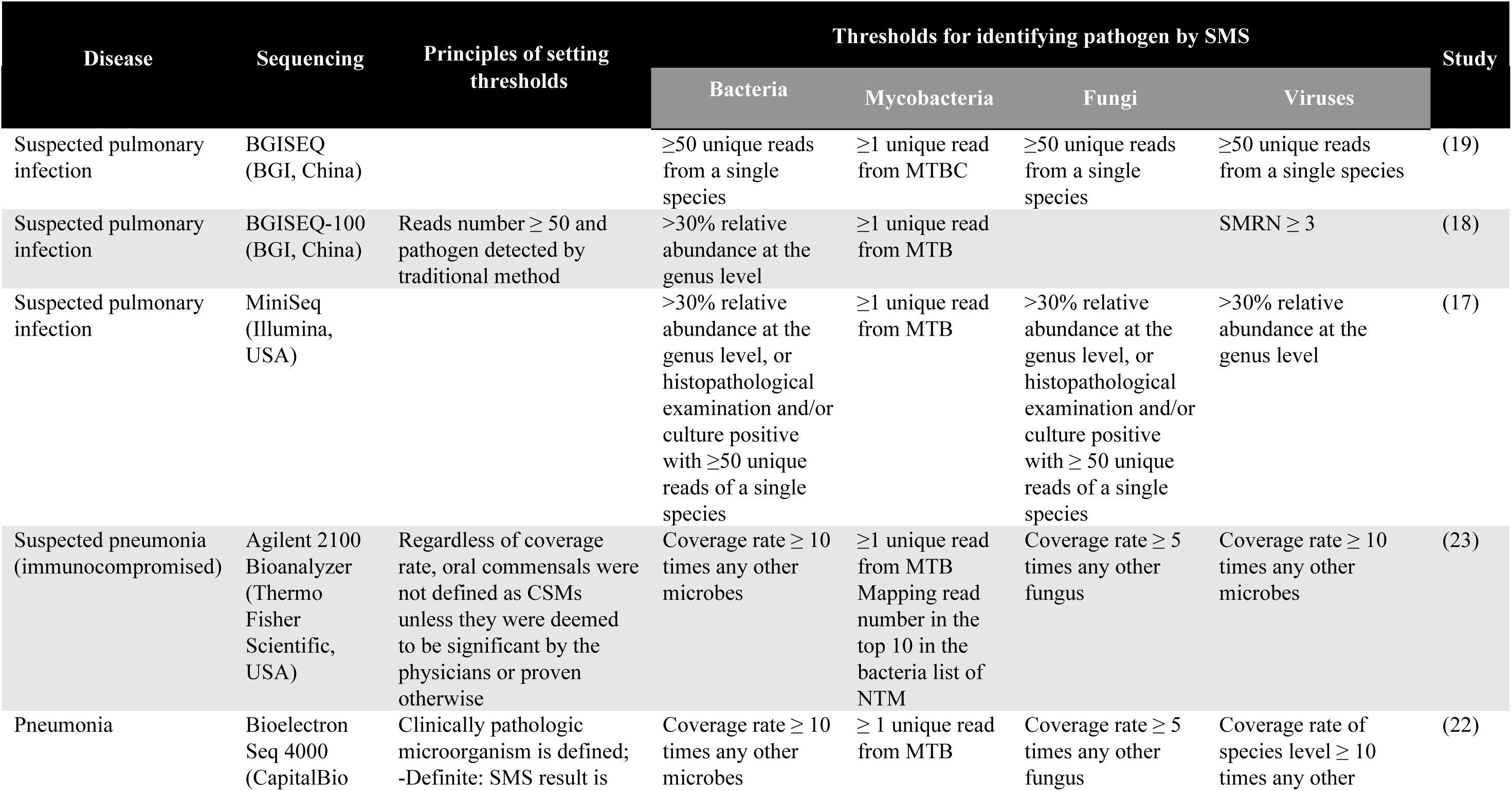
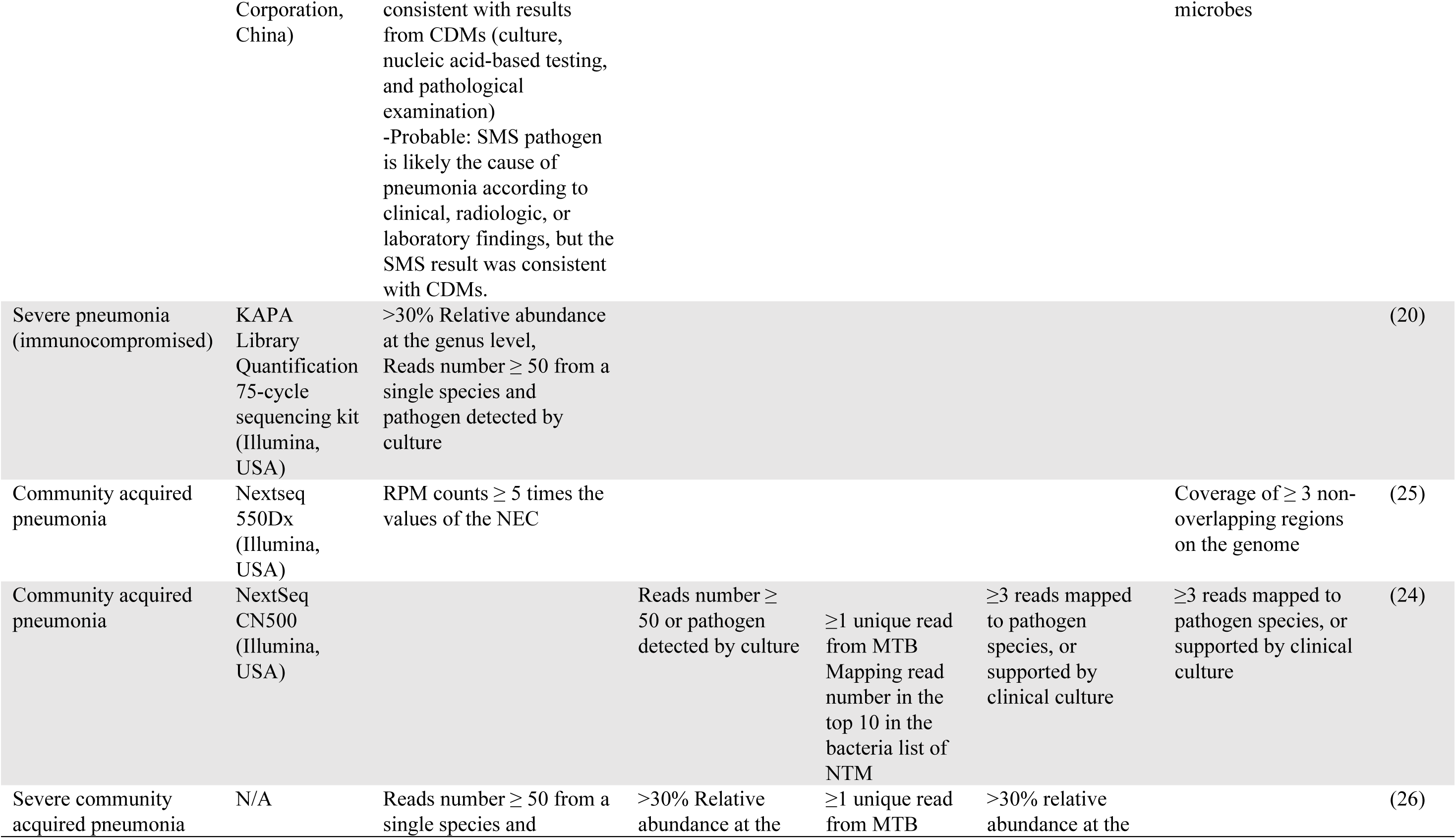
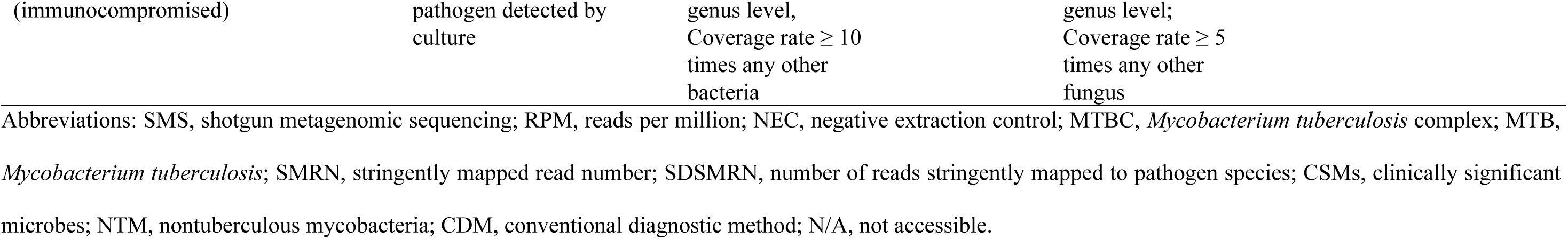
Thresholds for identifying clinically significant microbes using SMS.

Despite applying expanded criteria, SMS did not detect the pathogen in 31% of cases (5 out of 16). This included four cases (involving MTB, *Aspergillus* sp., or CMV) in which no reads were detected, and one case in which FA-PP identified *K. pneumoniae* at 10^5^ copies/mL; however, no corresponding reads were found in SMS.

Although MTB was detected in cases 1 and 2 by CDMs, SMS did not identify any MTB reads. This contrasts with previous studies that report a sensitivity of 44–48% for MTB detection by SMS, comparable to PCR (36, 37). Some studies have reported higher SMS sensitivity than target-specific real-time PCR (38, 39); however, these studies included diverse specimen types, such as lung biopsy tissue, limiting direct comparisons. Several factors may account for the lower sensitivity observed in this study. First, the nucleic acid extraction method used for SMS may have been suboptimal for mycobacteria. A study comparing sputum DNA extraction kits for *Mycobacterium* spp. demonstrated significant differences in 16S rRNA gene cycle threshold (Ct) values depending on the kit used (40). While no direct comparison exists between the QIAamp DNA Mini Kit used in this study and those used in previous SMS studies, kit-related variations may have influenced the results. The robust, waxy cell wall of *Mycobacterium* spp. makes lysis challenging, often leading to low DNA yield (41). Additionally, a study on sputum lysis methods for *Mycobacterium* spp. reported differences in Ct value standard deviation based on lysis temperature, further emphasizing the impact of extraction conditions (42). Second, differences in sample preprocessing may have affected SMS sensitivity. Variations in centrifugal force and duration have been shown to influence *Mycobacterium* sedimentation, affecting recovery rates and smear sensitivity (43). Notably, our study applied a higher relative centrifugal force than previous studies that reported greater SMS sensitivity for MTB. Excessive centrifugal force can generate heat, potentially leading to bacterial injury and reduced DNA recovery (37, 38, 43). The absence of phenol treatment, which has been suggested to enhance DNA purity, may have further contributed to reduced detection (41, 44). Finally, the patients’ treatment history may have influenced SMS sensitivity. Previous studies have reported a significant decrease in MTB detection sensitivity from 76% in pretreatment samples to 31% in post-treatment samples (37). Both cases 1 and 2 involved patients with a history of anti-tuberculosis treatment, and neither case exhibited rifampin resistance. In case 2, *M. tuberculosis* was detected by the Xpert MTB/RIF assay and cultured; however, *Corynebacterium striatum* was the predominant isolate, suggesting that competition with other organisms and prior treatment may have reduced SMS sensitivity. Enhancing MTB detection may require developing targeted panels optimized for *Mycobacterium* specimen processing.

In case 6, in which *Aspergillus* sp. was detected by CDMs, SMS did not yield any reads, likely due to the incomplete or variable nature of ITS reference sequences, which complicates fungal identification via SMS (45). In this study, fungal species identified via CDMs were not detected as positive reads using SMS. The ITS1 sequence is recognized for its reliability in identifying *Candida* spp., *Pneumocystis* spp., and *Aspergillus* spp. within the *Ascomycota* division, offering accurate species- and genus-level classification (46). Although fungal pneumonia is increasingly prevalent among immunocompromised individuals, all cases in this study were HIV-negative (13). Notably, *P. jirovecii* pneumonia detection has been increasing among HIV-uninfected immunocompromised individuals, highlighting its clinical relevance (47). However, SMS did not detect *P. jirovecii* reads in this study, even with the complete ITS1 sequence. Previous studies have reported significantly lower quantities of *Pneumocystis* spp. in BAL fluid from HIV-negative patients than from HIV-positive patients, which may have contributed to the challenges in SMS-based detection (48).

For cases 5, 6, and 16, CMV was considered the primary pathogen based on clinical assessment, with PCR results ranging from 9,665 to 200,175 copies/mL. In case 4, CMV was detected at 1,343,600 copies/mL; however, it was not targeted for treatment owing to the patient’s immunological status. These findings highlight the limitations of relying solely on CDMs for clinical decision-making, as CMV PCR results require integration with broader clinical context. Notably, SMS did not detect viral reads in any case, precluding the identification of CMV or other viruses. Despite DNA extraction yields exceeding 3–8 times the 4 nM threshold for library preparation, the proportion of valid viral sequences remained low (∼0.05%), indicating potential biases (49). The small size of viral genomes further complicates SMS-based detection (50). Although the DNA integrity number exceeded the manufacturer’s recommended threshold of 3, no standardized criteria exist for library construction. Additionally, while the Q30 score surpassed the 85% threshold, it did not reach 90%, which may have affected viral read recovery (51). Moreover, extended storage and delayed processing for SMS, compared to immediate testing for FA-PP and CMV PCR, may have contributed to viral degradation and reduced detection rates (52). Bacteriophages, which constitute the majority of viral particles in both environmental and human-associated microbiomes, account for over 90% of the human virome (53–55). Incorporating bacteriophage detection strategies into viral metagenomic analyses may enhance SMS-based viral identification.

All bacteria detected as positive by SMS in this study were gram-negative. Gram-negative bacilli are the most common cause of LRIs in elderly patients (4), and the 10 Gb SMS demonstrated strong performance in detecting these pathogens. Unlike previous studies, this study incorporated FA-PP into CDMs, potentially enhancing the pathogen detection rate. FA-PP has been shown to identify target pathogens even in samples reported as "no growth" or "normal flora" in culture (56). However, no clear correlation was observed between FA-PP semi-quantitative values and SMS read numbers. For FA-PP values of 10^5^ copies/mL, SMS reads ranged from 5–52; for 10^6^ copies/mL, reads ranged from 2–310; and for values exceeding 10^7^ copies/mL, reads ranged from 347–23,869. A culture result of 10^4^ CFU/mL corresponded to 10^5^ copies/mL in FA-PP, 3,000–6,000 CFU/mL corresponded to 10^6^ copies/mL, and ≥10^5^ CFU/mL aligned with 10^6^–10^7^ copies/mL in FA-PP.

This study partially explored genotypic AST using SMS in BAL fluid; however, results did not consistently align with those of phenotypic AST in cultured colonies. Using the CARD database (34), SMS-detected ARGs were compared with phenotypic AST results from CDM-identified pathogens; however, ARGs could not be definitively linked to the cultured strain. SMS also lacks the ability to trace specific ARGs to their bacterial origin (57), which may overestimate its predictive capacity, particularly when resistance is limited to a single antibiotic within a class. Although ARGs have been identified at the DNA level, their functional expression at the RNA or protein level remains unknown, limiting insights into actual resistance mechanisms. A meta-analysis of genotypic AST using metagenomic sequencing reported a categorical agreement of 88%; however, very major errors (VME) (24%, 95% CI: 8–40%) and major errors (ME) (5%, 95% CI: 0–12%) exceeded the US Food and Drug Administration (FDA)-recommended thresholds (57). While a machine learning-based genotypic AST model met FDA requirements (ME ≤3%, VME ≤1.5%) with high performance (58), clinical specimens pose additional challenges. The presence of host nucleic acids, multiple pathogens, and plasmid-mediated ARGs complicates species attribution (58–60). Additionally, database limitations and low area under the curve (AUC) values in some models highlight the need for further research to refine genotypic AST approaches (58).

However, the direct application of SMS-detected ARGs in clinical practice remains premature. While SMS cannot conclusively determine the direct involvement of ARGs in resistance phenotypes, it enables the detection of microbes at very low concentrations and provides a comprehensive overview of the resistome within a sample (56, 57). The administration of antibiotics to patients colonized with multidrug-resistant organisms increases colonization density and expands the resistance gene pool, thereby increasing the risk of infection (58, 61). The maintenance of resistance-conferring mutations in bacterial pathogens is entirely dependent on their effect on fitness and virulence (62). Given the association between the resistome and microbial diversity, antimicrobial stewardship is increasingly emphasized. Overcoming current limitations may allow SMS-based ARG identification to contribute to these efforts (58).

This study demonstrates the feasibility of achieving sufficient sequencing coverage for SMS with a 10 Gb output. Previous studies have not provided detailed sequencing configurations for BAL fluid. While highly complex samples such as stool require 1–10 Gb (with >7 Gb recommended), lower-complexity samples such as anterior nares may be adequately analyzed with <1 Gb (63, 64). To assess whether 10 Gb was sufficient for BAL fluid analysis, we conducted experiments based on existing recommendations (35). Increasing sequencing output generally enhances pathogen detection; however, in untargeted approaches without host genome depletion, a proportional increase in host-derived sequences may limit improvements in sensitivity (65). Using a 10 Gb configuration, DNA-based SMS detected primary pathogens in 63% of cases based on applied thresholds, increasing to 69% when subdominant taxa were included. In SMS, the presence of host nucleic acid affects sensitivity, and appropriate depletion can increase the relative abundance of microbial signals (66). However, unintentional removal of microbial content along with host DNA and the variability of results depending on sample type remains controversial (67). In this study, SMS was conducted without host genome depletion. Further research is required to validate the effective host signal removal in BAL samples.

Inconsistencies between SMS and clinical diagnoses are well-documented, with comparative studies reporting low agreement levels (κ = 0.035–0.347) between SMS and CDM positivity (22, 23, 26). In this study, cases with negative SMS results despite positive CDM findings underscore the limitations of SMS as a molecular diagnostic tool (68). Challenges include differentiating viable pathogens from residual nucleic acids in resolved or treated infections and distinguishing asymptomatic colonization from true infection (14). Additionally, variations in bacterial read distributions complicate the application of uniform positivity thresholds. Future studies should refine SMS interpretation criteria by integrating clinical features to improve their correlation with clinical outcomes. Comparative analyses of sequencing output capacities for BAL fluid samples are required to determine the optimal conditions for pathogen detection. Further research should also explore methodologies that expand SMS applications beyond the bacteriome to include the mycobiome and virome, thereby enhancing its diagnostic potential.

## Conclusions

This study presents a comparison between SMS and CDMs, elucidating the performance and limitations of SMS with a 10 Gb output for LRI diagnosis. While SMS cannot replace PCR, it can complement culture methods, serving a mutually supportive role. To establish SMS as a valuable supplementary tool in LRI diagnostics, further research is needed to explore its variable applications for higher sensitivity and cost-effectiveness.

## Data Availability

The raw sequencing data were submitted to the NCBI Sequence Read Archive (SRA) database (accession number PRJNA-1036216).

## Acknowledgments

We express our profound gratitude to the laboratory and medical personnel for their diligent execution of clinical measurements, whose contributions have been instrumental in facilitating the success of this project. We extend our appreciation to the reviewers for their valuable suggestions and insights. We convey our gratitude to all the authors for their dedicated efforts toward the culmination of this project.

## Author contributions

**Conceptualization:** Ha-eun Cho and Young Jin Kim**. Data curation:** Ha-eun Cho, Min Jin Kim, Jae Joon Lee, Sun Young Cho and Kyung Sun Park. **Formal analysis:** Ha-eun Cho, Ki-Ho Park and Young Jin Kim. **Investigation:** Ha-eun Cho and Min Jin Kim. **Methodology:** Min Jin Kim, Jongmun Choi and Yong-Hak Sohn. **Supervision:** Young Jin Kim. **Writing – original draft:** Ha-eun Cho. **Writing – review & editing:** Young Jin Kim.

